# LEVEL OF FOOD CONSUMPTION SCORE AND ASSOCIATED FACTORS AMONG PREGNANT WOMEN AT SHEGAW MOTTA HOSPITAL, NORTHWEST ETHIOPIA

**DOI:** 10.1101/2020.04.22.20076034

**Authors:** Mehariw Birhan, Tsegahun Worku, Mekuanint Taddele, Zewdie Aderaw

## Abstract

**Background:** All populations are at risk for poor food consumption, but pregnant women are the most vulnerable groups for poor food consumption particularly for those in resource limiting settings like Ethiopia. However, there is a lack of literature for food consumption score in these population groups and study area. Therefore, the results of this study may give information for local decision makers.

**Objective:** To assess level of food consumption score and associated factors among pregnant women attending antenatal service at Shegaw Motta Hospital, Northwest Ethiopia, 2018.

**Method:** Institution based cross-sectional study design was conducted among 422 pregnant women attending antenatal care service at Shegaw Motta Hospital from February 23, 2018-April 3, 2018. A systematic random sampling technique was used to select study participants. Data were collected using interviewer administered structured questionnaire and coded and entered to Epi-Data version 3.1 then exported to SPSS version 23 for analysis. Binary Logistic Regression was used for analysis.

**Result:** A total of 422 pregnant women with response a rate of 100% participated in the study. The overall food consumption score among pregnant women was found to be eight of study participants [1.9% (95% CI: 0.7, 3.3)] reported that their food consumption score were poor, seventy of them [16.6% (95% CI: 13.0, 20.4)] were borderline and the remaining 81.5 % (95% CI: 77.5, 85.1) of them had acceptable food consumption score. Residence, being rural or urban [AOR=4.594(95% CI: 1.871, 11.283)], religion status, being an Orthodox or others [AOR= 0.073(95% CI: 0.021, 0.254)], were factors associated with food consumption score.

**Conclusion and recommendation:** Food consumption scores among pregnant women was found to be highly unacceptable. Residence and religion were factors associated with food consumption score. Therefore, appropriate nutrition education should be given.

## 1. INTRODUCTION

### 1.1. Background

Food consumption score (FCS) is a composite score based on dietary diversity, food frequency and relative nutritional importance of different food groups. Information is collected from a country specific list of food items and food groups over the recall period of the past seven days, then, consumption frequency of each food group is multiplied by assigning weight based on its nutrient content and finally the value are summed to obtain the food consumption score (1-2).

All human beings need a balanced amount of nutrients for proper functioning of the body system because nutrition is a fundamental pillar of human life, health and development throughout the entire life(3). The index of diversity and balanced food consumption is called Food Consumption Score (FCS), is the core indicator of consumption. It has been used in several report or analysis around the world apart from being the selected indicator for the most recent analysis of the food security in Yemen (1). The score is calculated using weighted frequency of consumption of nine food groups consumed by a household during the seven days before the study. The weights for each group of food are listed in the world food program-vulnerability assessment and monitoring (WFP-VAM) and are based on the nutrient density analysis. Higher or lower weights are assigned in terms of caloric density and macro and micro nutrient content (1, 4).

Having all this information, Food Consumption Score is calculated according to the following ways: sum all the consumption frequencies of food items of the same group, and recoded the value of each group above 7 as 7 then multiply the value obtained for each food group by its weight and sum of the weighted food groups scores, thus creating the Food Consumption Score (FCS); as follows <21.5 poor food consumption, 21.5-35 borderline food consumption and >35accepyable food consumption. Scoring below 28 is poor food consumption; if a household has a diet that does not include at least a staple, oil, sugar, and vegetables on a daily basis (1-2).

### 1.2. Statement of the Problem

Several studies show that the nutrition problem is still highly prevalent especially in pregnant women and children (5). Maternal under-nutrition diminishes woman productivity, causing bad results for herself, her family, her community, and the broader society. Maternal malnutrition is influenced not only by lack of adequate nutrition but also influenced by factors like socio demographic factors, nutritional knowledge of mother during pregnancies (6).

The prevalence of food consumption score with poor and borderline food consumption provides essential information on people’s current diets and is helpful in deciding the most appropriate type and scale of food security intervention as well as the right target group like pregnant women and children for the assistance. Having such information could enhance inputs into nutrition-sensitive program design as well as improving measurement of food-based interventions (4, 7). However, the current financial crisis seems to have affected consumers’ attitudes in many countries. The economic uncertainty and insecurity have led consumers to take decisions minimizing their costs, even for basic needs, such as food quantity and quality. In a period of inflation and unemployment, consumers are more likely to change the composition of their expenditures therefore there is a strong shift towards poor food consumption for cheaper products with lower nutritional value that leads malnutrition (7).

Several food security studies show that many households, especially live in the rural areas, are food insecure. The problem is not only about food shortages but inadequate access of food by vulnerable groups (8-9).

Government gives much attention to the subsector to increase production and marketing through accelerated investment in infrastructure and adoption of better seed varieties and fertilizer technology to assure food consumption(10). In particular, teff, wheat, corn, maize and sorghum are the most staple food items in the study area that are monotonous which leads to poor food consumption (11). Due to this poor food consumption both chronic and acute problems of food consumption are widespread and severe in both rural and urban areas of the country (12).

Even though, some study conducted about food insecurity which is a major indicator of food consumption score has given more emphasis to the rural area of the country do not identify situations at grass root level (7, 13).

Therefore, the research undertaking at Shegaw Motta Hospital is essential to identify level of food consumption score and factors among pregnant women since the results may give information to develop and inter in to nutrition intervention activities for Shegaw Motta hospital, Huleteju Woreda Health Office, Agricultural office, Educational office and District administrational office in order to minimize its problem.

### 1.4. Justification of the study

Chronic energy deficiency is caused by eating too little or having an unbalanced diet that lacks adequate nutrients which is a major risk factor for adverse birth outcomes. Unacceptable food consumption score (poor or borderline) is the major public health problem when the score is greater than ten which is called catastrophe that highlighting nutrient inadequacy which is associated to food insecurity, under nutrition, growth failure, reproductive failure, anemia, malaria, diarrhea, unfavorable pregnancy outcome (abortion, stillbirth) and vitamin A deficiency. Despite there is an improvement in the implementation of nutrition intervention since 2008 in Ethiopia, unacceptable food consumption score 26% in nationwide and 11% Amhara region that did not show significant reduction so the government has given attention by designing different programs like National Nutrition Program II (NNP II) which is working to address under nutrition in the first 1000 days from conception up to 2 years children’s lives can be saved and can grow healthily and fullest potential throughout the life. East Gojjam Zone including the study area is highly food secured, but under nutrition is the major problem.

The food consumption score is a catalyst towards improved nutrition sensitive and specific programs by identifying nutrient inadequacies in households, but the relationship between food consumption score and associated factors are not done in Ethiopia to address the problem. It can also have an indirect impact on the lives of many more people by advocating for comprehensive solutions and supporting local decision makers to develop strategies to overcome under nutrition. There is limited information in Ethiopia in general and particularly in the study area about the level of food consumption score and associated factors.

The finding of this study will give information for Shegaw Motta Hospital, Woreda health office, Town health office, Education office, and Agricultural office to develop strategies that could optimize nutrition intervention activities among pregnant women. It may also give clues for researchers to conduct further researches.

## 2. OBJECTIVE OF THE STUDY

### 2.1. General Objective

To assess the level of food consumption score and associated factors among pregnant women attending antenatal care service at Shegaw Motta Hospital, Northwest Ethiopia, 2018

### 2.2. Specific Objectives

1. To determine the level of food consumption score among pregnant women attending antenatal care service
2. To identify factors associated with food consumption score among pregnant women attending antenatal care service

## 3. METHODS AND MATERIALS

### 3.1. Study Design

An institution based cross-sectional study design was used.

### 3.2. Study Area and Period

The study was conducted at Shegaw Motta Hospital. Shegaw Motta Hospital is located in East Gojjam Zone, Northwest Ethiopia, which is far from 372 Km from Addis Ababa to Northwest direction, 120 Km from Bahir Dar to south direction and 200 Km from Debre Markos, this Hospital has a latitude and longitude of 11°5’N 37° 52’E and 11.083°N 37.867°E respectively with an elevation of 2,487 meters (8159 feet) above sea level. The weather condition is categorized in Woyna Dega and Shegaw Motta Hospital has more than 500,000 catchment population. Regarding the dietary practice Teff, Wheat, Sorghum, Maize, Bean, Chickpea, Pea and other cereals and legumes are the common food sources whereas; fish and other seaweeds are not easy to access. The obstetric and gynecological care has started at the beginning of Shegaw Motta Hospital establishment 18 years ago. Currently the hospital is providing antenatal care for more than 9,900 pregnant women per year and 1050 pregnant women were enrolled in one month and get the service with 17 health care providers that is 14 midwives, 1 gynecologist and 2 emergency surgery officers (11). The study was conducted from February 23, 2018-April 3, 2018.

### 3.3. Population

#### 3.3.1. Source Population

All pregnant women attending antenatal care service at Shegaw Motta Hospital.

#### 3.3.2. Study Population

All pregnant women attending antenatal care service at Shegaw Motta Hospital during the data collection period.

### 3.4. Eligibility Criteria

#### 3.4.1. Inclusion Criteria

All pregnant women who were attended antenatal care service at a Shegaw Motta Hospital and consumed only home prepared foods in the last seven days during the data collection period.

#### 3.4.2. Exclusion Criteria

Pregnant women who consumed any food item which was not prepared at home were excluded.

### 3.5. Sample Size Determination and Procedure

#### 3.5.1. Sample Size Determination

For the first objective, the sample size was computed based on a single population proportion formula assuming the prevalence (p) of food consumption score 50% (because there was no previous study conducted in Ethiopia), 95% Confidence level (1.96), and 5% margin of error.

n= [(Z□/2)^2^ *P (1-P)] **/**d^2^

Where;

✓ n = estimated sample size
✓ Zα/2 =a standard normal value which corresponds 95% of confidence level = 1.96
✓ p= estimated prevalence of the food consumption score is (0.5).
✓ d= tolerable sampling error = 0.05

After substitution the estimated sample size = 384,

Taking 10% of non response rate, the final sample size was **422**.

Sample size for the second objective was determined using double population proportion formula by Epidemiology Information (Epi info) statistical calculation considering 80% power and one to one ratio based on a study conducted in Gondar town, Northwest Ethiopia, 2014 (14).

#### 3.5.2. Sampling Procedure

Systematic random sampling technique was employed to select study participants. According to Shegaw Motta Hospital ANC Report Sheet, in average a total of 1,050 pregnant women attends ANC per month. Therefore, 422 study participants were selected by systematic random sampling technique. By taking the final sample size (n= 422), K was two. Thus, the study participants were selected every 2^nd^ interval. To get the initial study participant, lottery method was used. Then, each study participant was selected every 2^nd^ interval using systematic random sampling technique. But, when the selected study participant did not fulfill the inclusion criteria, the next individual was included.

### 3.6. Variables of the Study

#### 3.6.1. Dependent Variable

Food consumption score

#### 3.6.2. Independent Variables

##### Socio-demographic variables

Age, religion, occupation, residency, educational status, marital status, husband employment, and husband educational status.

##### Obstetric factors

Gestational age, parity, gravidity, number of ANC follow up

##### Knowledge

of food consumption score

##### Attitude

for food consumption score

**Wealth index**

#### 3.7. Operational Definition

##### Unacceptable food consumption

The respondent has not eaten Base diet at least staples, fruit, vegetables and oil <= 3days per week is called unacceptable food consumption score(2, 15).

##### Acceptable food consumption score

Base diet should be >=36 (>=4 days per week of daily base food) absence of other food groups truly acceptable diet (2, 15).

##### Knowledge

is awareness and understanding that pregnant women have gained on food consumption during pregnancy through learning and practice.

##### Adequate Knowledge

pregnant women considered being knowledgeable if she correctly answered greater than or equal to 14 of the total 19 knowledge related questions (16).

##### Inadequate knowledge

pregnant women considered to have inadequate knowledge if she has answered less than 14 out of the total 19 knowledge related questions (16).

##### Attitude

feeling or perception that pregnant women have gained towards food consumption.

##### Favorable attitude

Participants who answered greater than or equal to 15 of the total 20 attitudes relating questions.

##### Unfavorable attitude

Participants who answered less than 15 out of the total 20 attitude questions.

##### Wealth index

The composite indicator of socioeconomic status, which was computed by the application of principal component analysis (PCA). Initially household asset data were prepared for analysis. Before the PCA, using frequency, important variables that can discriminate households were selected to reduce the number of variables. The binary variables were coded to 0 and 1and categorical variable options were converted into binary variables and dummy variable was created as 0 and 1.After data preparation, variables were standardized to change variables in to the same scale for comparison by subtracting the mean from each value and then dividing the standard deviation. Once standardized, the variables have mean of zero and standard deviation of 1.

A total of 17 variables was considered for wealth index construction. However, 12 variables were dropped as their communality scores were less than 50%. The rest five variables, including number of cattles in the household, number of rooms in house, having commercial bank account, presence of agricultural land including irrigation and type of latrine to the household were considered for wealth index construction.

In PCA, the sum of components with Eigen values greater than one should explain at least 60% of the total variance (17).In this study, the components explained 79.6% of the total variance, which was above the recommended minimum value (17).Wealth index values were calculated by summing up the scores for the five components. Finally, the index was developed by categorizing the sum of components into five equal parts and the parts were ranked from the poorest to the wealthiest.

### 3.8. Data Collection Procedure and Tool

Data were collected using interviewer-administered structured questionnaire which was developed based on World Food Program (WFP) standardized eight food frequency questions, Ethiopian Demographic and Health Survey and different literatures. The tool contains socio-demographic characteristics, obstetric, attitude, knowledge, household assets and food consumption questions. There is standardized World Food Program (WFP) eight food groups English version questionnaire and this questionnaire was translated to Amharic language. The respondents asked to recall all foods and beverages they have taken in last seven days prior to the interview to assess food consumption score of the respondents. Using Ethiopian Food Composition Table the local food items were categorized into eight food groups. All consumption frequency of foods in the same group were summed and multiplied by the value of each food group by its weight. The details are described in **Table 2**. Then the weight of food groups score were summed to obtain food composition score to determine the status of pregnant women food consumption score based on the following result: 0-21 score was poor, 21.5-35 score was borderline and **>**35score was acceptable. In general if food consumption score is <35 it indicates the presence of household food insecurity (1).

**Table 1:**
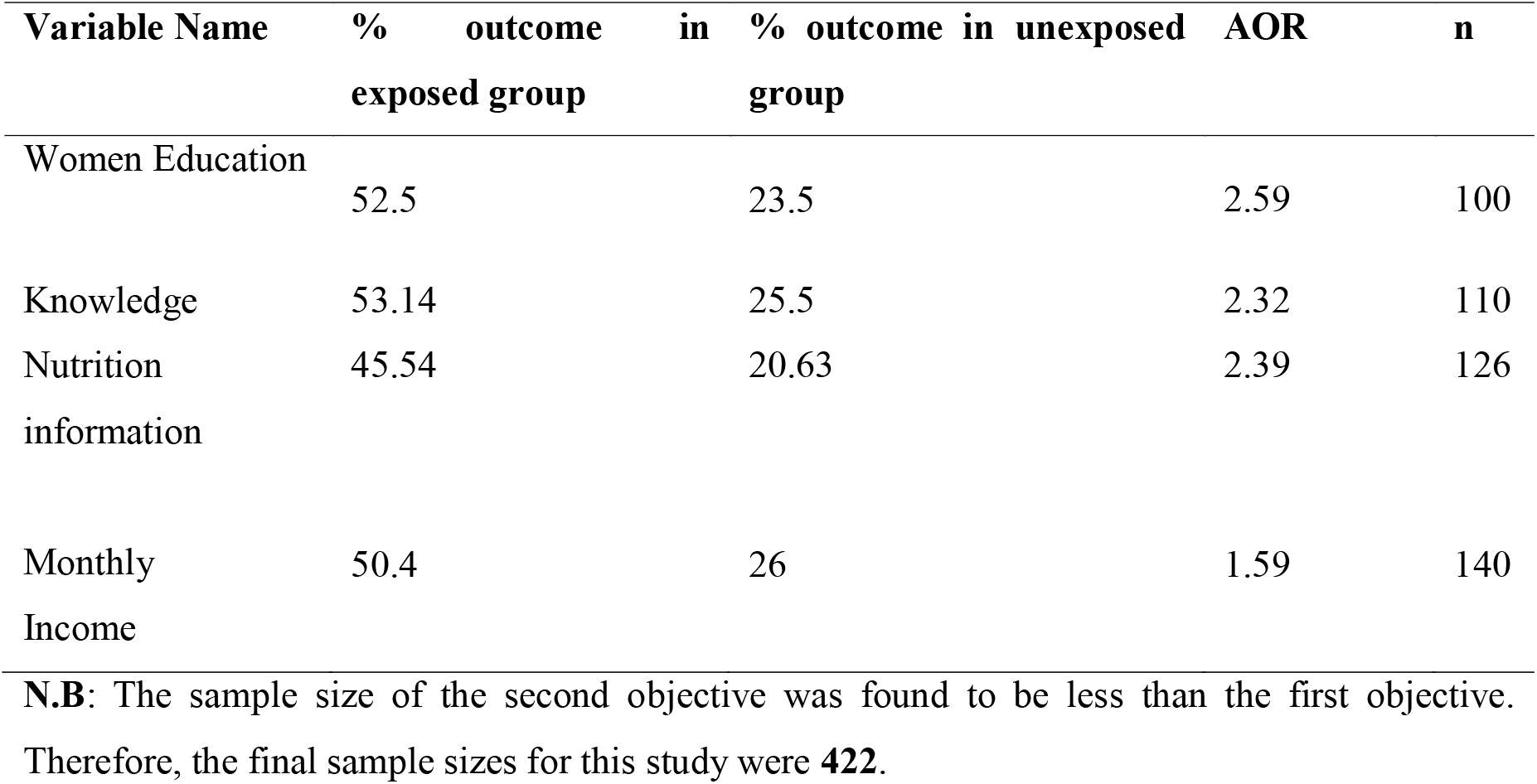
Sample size calculation of the second objective for a study conducted among pregnant women food consumption score at Shegaw Motta Hospitals, February 23, 2018–April 3, 2018.

**Table 2:**
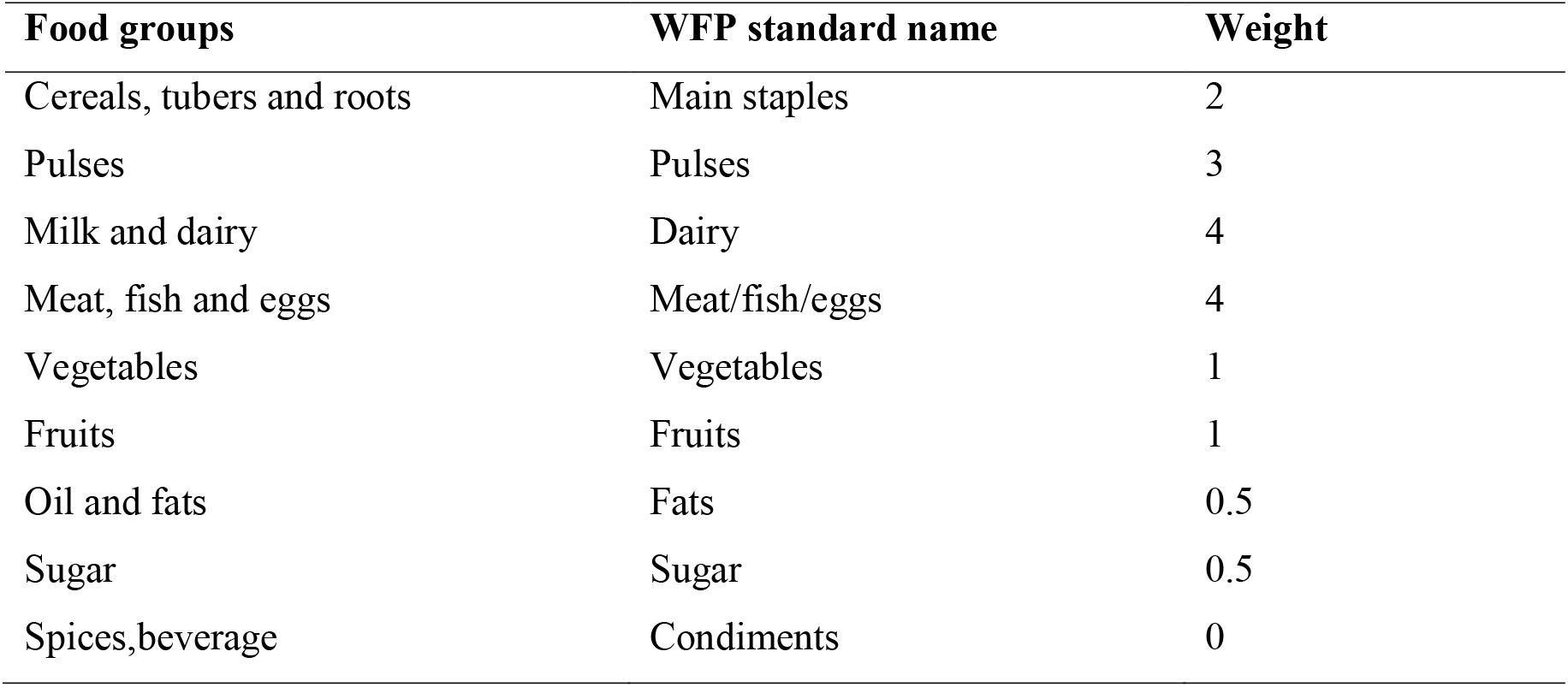
World Food program (WFP) eight food groups with each standard weight, 2015 G.C.

| **Food groups** | **WFP standard name** | **Weight** |
| --- | --- | --- |
| Cereals, tubers and roots | Main staples | 2 |
| Pulses | Pulses | 3 |
| Milk and dairy | Dairy | 4 |
| Meat, fish and eggs | Meat/fish/eggs | 4 |
| Vegetables | Vegetables | 1 |
| Fruits | Fruits | 1 |
| Oil and fats | Fats | 0.5 |
| Sugar | Sugar | 0.5 |
| Spices, beverage | Condiments | 0 |

Two professional midwives (data collectors) and one Gynecologist (supervisor) were recruited to collect the data. The supervisor was followed the data collection technique and all the data collectors and the supervisor had regular meetings with the principal investigator at the end of each day of data collection period.

### 3.9. Data Quality Assurance

The qualities of the data were maintained by translating the English version questionnaire to Amharic language, then it was translated back to English to check its consistency. Pretest was conducted on 5% of the calculated sample size at the Mertule Mariyam primary hospital and necessary amendment was made. Prior to data collection, data collectors and supervisor were trained in one day by the nutritionist about overall purposes of the research, how to approach the study participants, and the way how to collect data. Furthermore, the principal investigator was provided feedback on a daily basis to the data collectors. Completeness, accuracy, and clarity of the collected data were checked carefully on a regular basis. To minimize recall bias and social desirability bias, study participants were asked probing or leading question or were asked relating to the local events like the date of Saint Marry and isolation of the respondents respectively.

#### 3.10. Data Analysis Technique

The collected data were coded and entered into the Epi-Data Version 3.1 and cleaned. The cleaned data set was exported to SPSS Version 23.0 software for analysis. Study participants food consumption score (poor, borderline and accepted), socio-demographic characteristics and other independent variables were presented using relevant descriptive statistics. Univariate analysis was done the 25% level of significance to screen out potentially significant independent variables. Multiple Logistic Regression was performed to see the association between the dependent variable and independent variables. The adequacy of the final model was checked using Hosmer and Lemeshow goodness of fit test, and an assumption of Binary Logistic Regression such as no multico linearity was checked. For Binary Logistic Regression, 95% confidence interval was calculated and variables with p-value < 0.05 were considered as statistically significant with the outcome variable (food consumption score).

## 3. Ethical Consideration

The ethical clearance was obtained from Debre Markos University, college of health sciences, departments of Public Health. Official letter was written along with the ethical clearance and submitted to Shegaw Motta Hospital by the principal investigator. In addition, prior to data collection permission was obtained from the hospital. Autonomy of participants was respected. This study had no any negative consequence for study participants. Participants were also informed that participation is voluntary. Written consent was taken from the study participants after briefing them the objective of the study. The confidentiality of participants related data were maintained by avoiding possible identifiers such as name of the respondents,the only identification number was used as a reference. The interviewer administered questionnaires were kept safe throughout the whole process of the research work.

During the data collection time any woman with medical problem findings such as abortion, education was provided and contacted health professionals in Shegaw Motta hospital for support and treatment.

## 4. RESULTS

### 4.1. Socio-demographic characteristics

A total of 422 study participants was included in the study making a response rate of 100%. The mean (±Standard deviation) age of the study participants was 27.64(±5.426) years. All most all (99.5%) were Amhara and more than half of study participants, 58.1% were in the age range 25-34 years. More than two third (73.2%) of study participants live in urban areas. Vast majority of the pregnant women were married (99.1%) and orthodox Christian in religion (71.3%). Besides, 25.4% of pregnant women had attended diploma and above educational status and 37.6% of pregnant women were merchants. Concerning on husbands’ education and occupation, 34.7% were diploma and above, and 36.7% were merchants. Regarding wealth status of pregnant women, 114 (27%) of the study participants were poorer and 107 (25.4%) of them participants were rich **(Table 3)**.

**Table 3:**
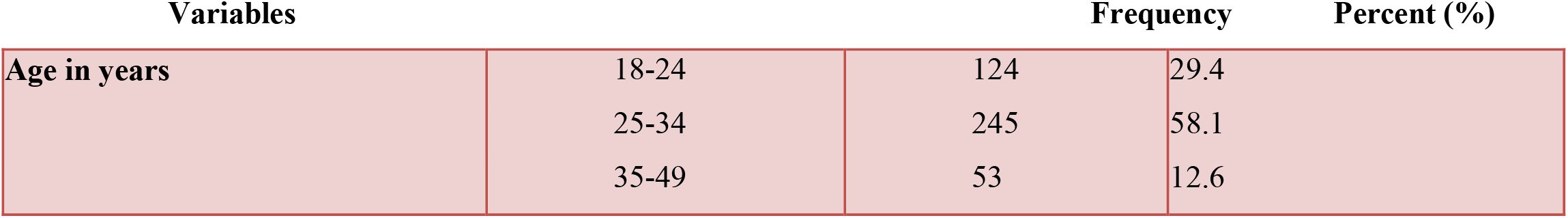
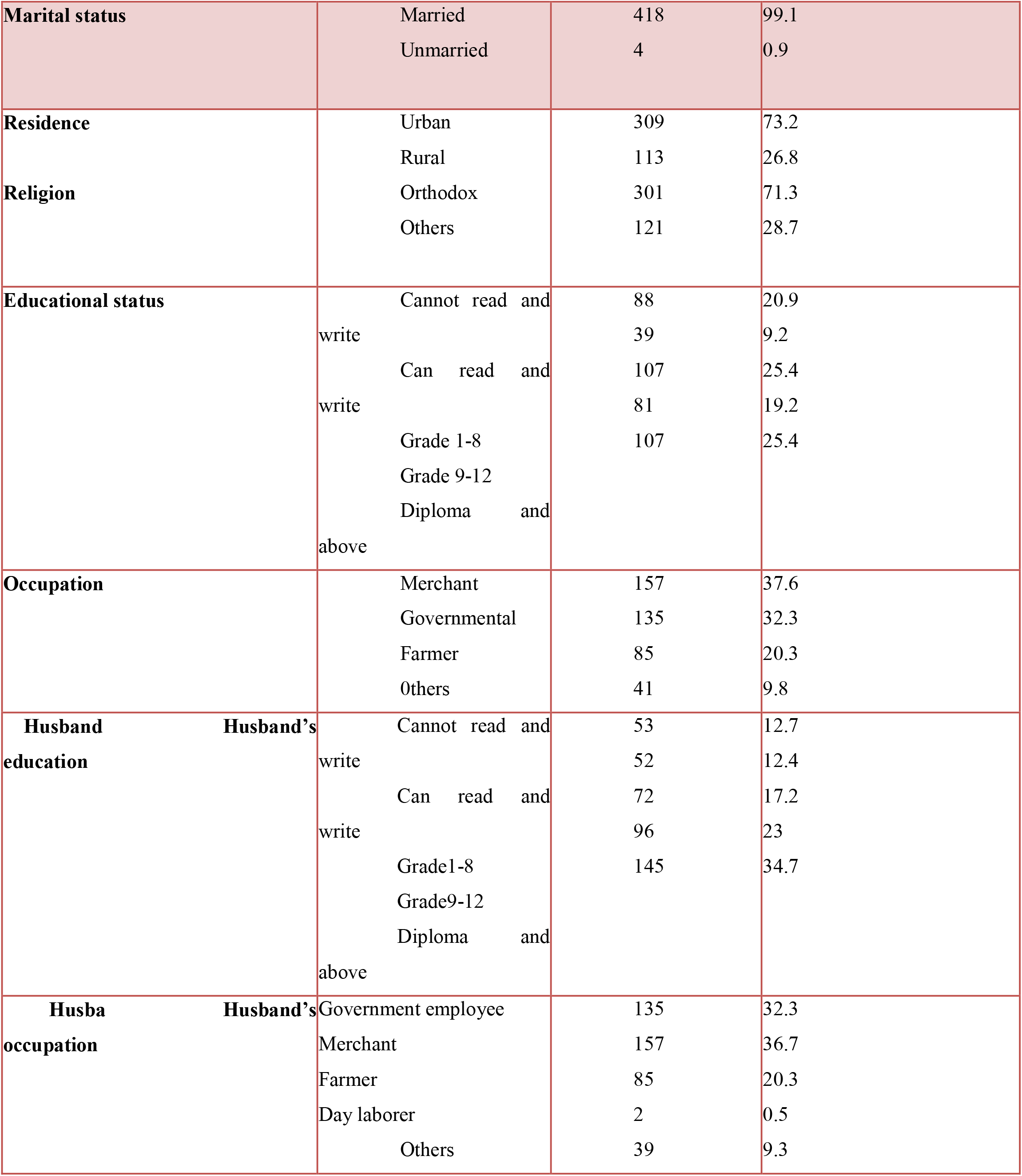
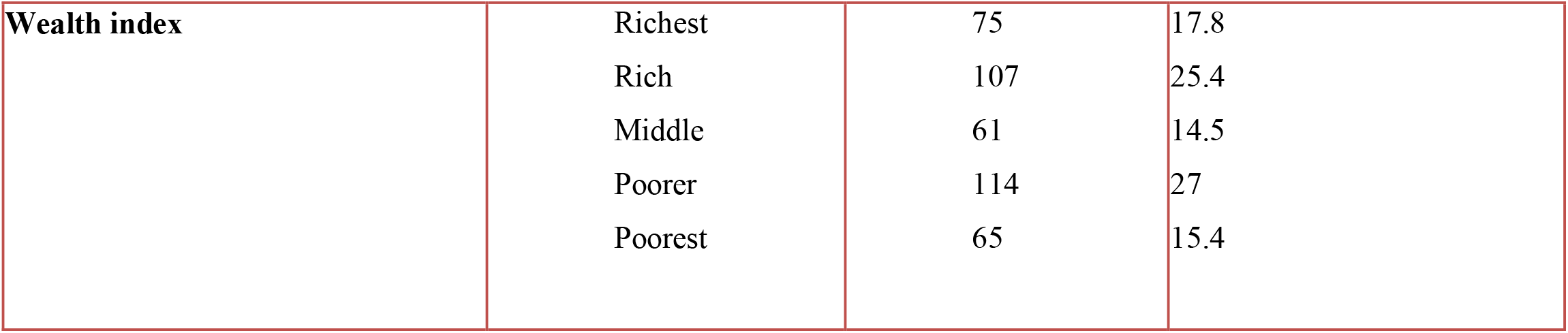
Socio-demographic characteristics of pregnant women attending the ANC service at Shegaw Motta Hospital, 2018 (n=422)

### 4.2. Food Consumption Score

Majority 344 (81.5%: 95% CI: 77.5, 85.1) of study participants’ acceptable food consumption score and around 70 (16.9 %:95%CI: 13.0, 20.4) and eight (1.9%: 95% CI: 07, 3.3) of study participants reported that borderline and poor respectively **(Table 4)**.

**Table 4:** Food Consumption Score of pregnant women attending the ANC service at Shegaw Motta Hospital, 2018 (n=422)

| **Variables** |  | **Frequency** | **Percent (%)** |
| --- | --- | --- | --- |
| **Food consumption scores** | Acceptable | 344 | 81.5 |
|  | Borderline | 70 | 16.6 |
|  | Poor | 8 | 1.9 |

### 4.3. Obstetric characteristics of study participants

Two hundred eighteen (51.7%) of study participants were at third trimester by gestational age and 282 (66.8%) of the study participants were multigravida. Concerning in parity, 171(40.5%) of the study participants had given birth at least once. About 346(82%) of participants were repeat ANC visitors. While 390(92.4%) and greater than three fourth (91.7%) of study participants did not have any history of stillbirth and abortion in their reproductive life respectively **(Table 5)**.

**Table 5:** Obstetric characteristics of pregnant women attending the ANC service at Shegaw Motta Hospital, Northwest Ethiopia, 2018 (n=422).

| Characteristics |  | Frequency | Percent |
| --- | --- | --- | --- |
| **Gestational age** | >=29weeks | 218 | 51.7 |
|  | 13-28weeks | 191 | 45.3 |
|  | <=12 weeks | 13 | 3.1 |
| **Gravidity** | Multigravida | 282 | 66.8 |
|  | Prim gravida | 140 | 33.2 |
| **Parity** |  |  |  |
|  | para one | 171 | 40.5 |
|  | Nulliparous | 141 | 33.4 |
|  | Multiparous | 110 | 26.1 |
| **Number of ANC visit** | >=2<sup>nd</sup> | 346 | 82 |
|  | 1<sup>st</sup> | 76 | 18 |
| **History of still birth** | No | 390 | 92.4 |
|  | yes | 32 | 7.6 |
| **History of abortion** | No | 387 | 91.7 |
|  | Yes | 35 | 8.3 |

### 4.4. Knowledge and Attitude of Food Consumption Score

Two hundred twenty four (53.1%) and 399 (94.5%) of the study participants had adequate knowledge about food consumption score and favorable attitude towards food consumption score respectively **(Table 6)**.

**Table 6:**
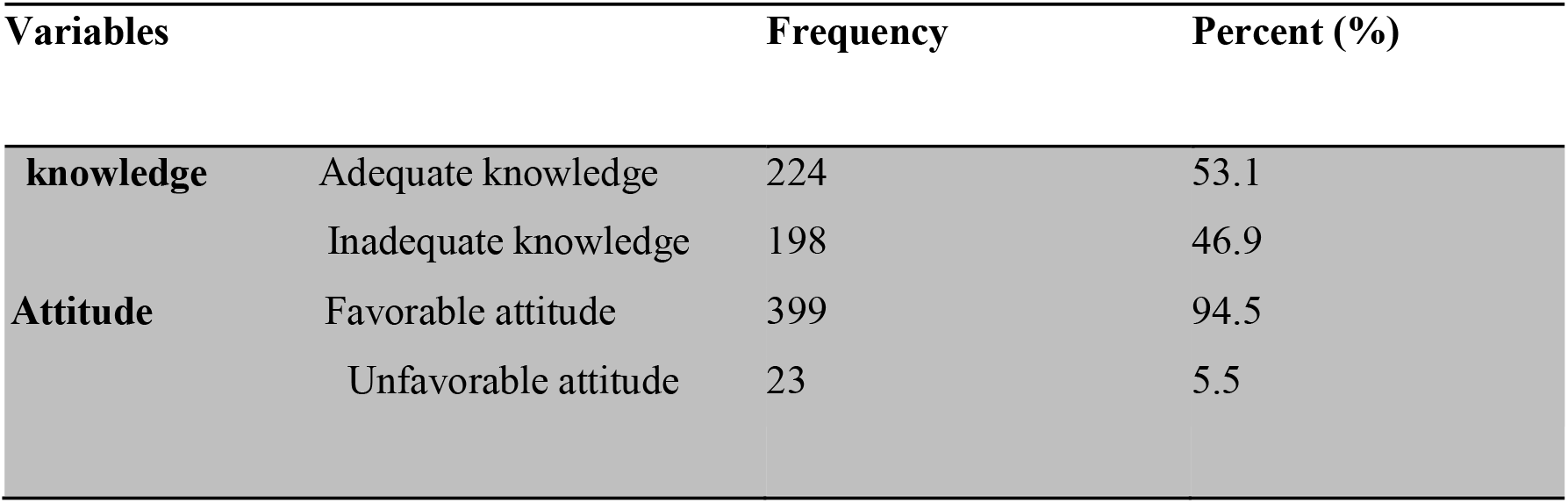
Knowledge and attitude of Food Consumption Score among pregnant women attending ANC service at Shegaw Motta hospital, Northwest Ethiopia, 2018 (n=422)

| Variables |  | Frequency | Percent (%) |
| --- | --- | --- | --- |
| **knowledge** | Adequate knowledge | 224 | 53.1 |
|  | Inadequate knowledge | 198 | 46.9 |
| **Attitude** | Favorable attitude | 399 | 94.5 |
|  | Unfavorable attitude | 23 | 5.5 |

**Table 7:**
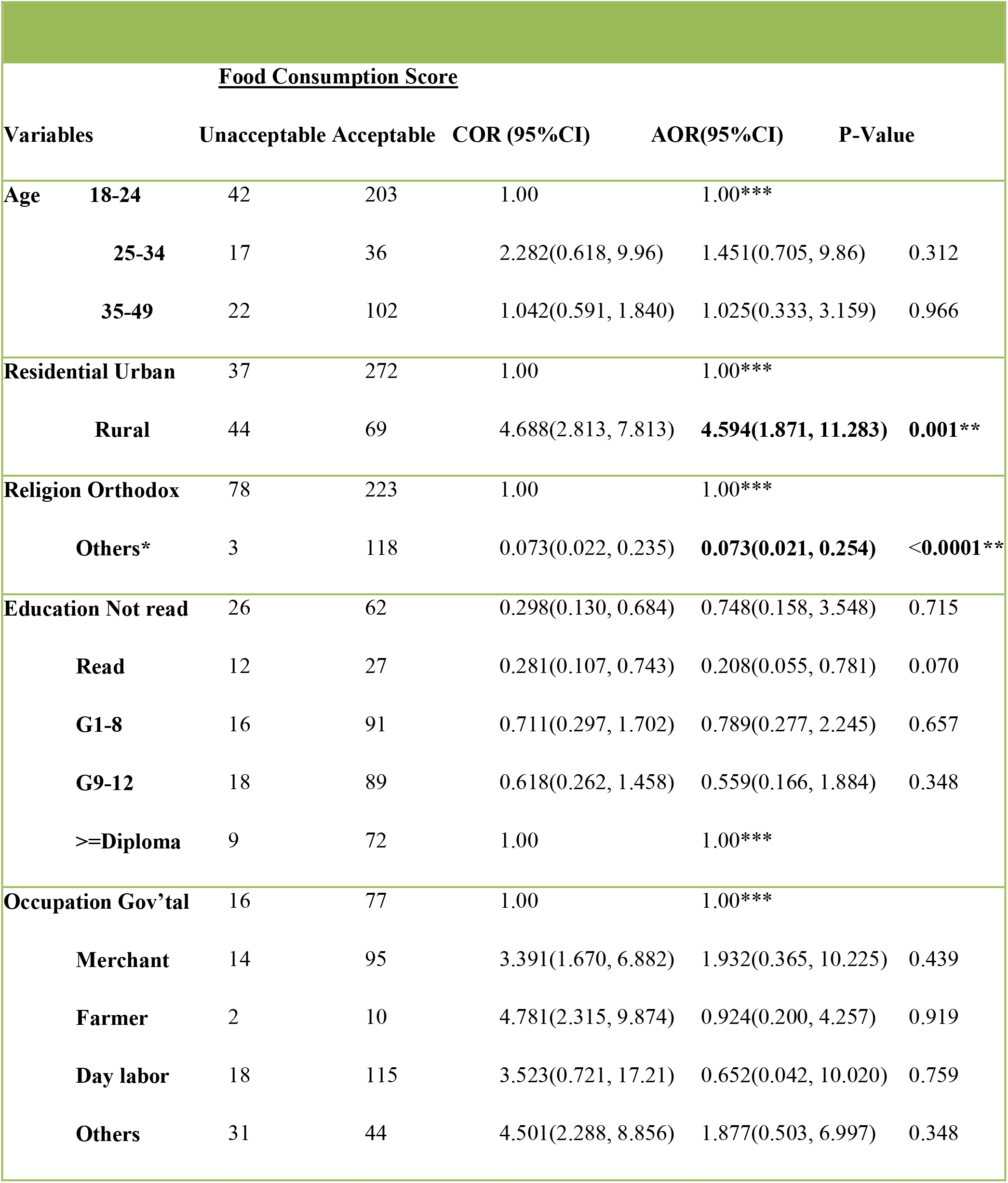
Univariate and Multivariable analysis for factors associated with food consumption score among pregnant women attending the ANC service at Shegaw Motta Hospital, Northwest Ethiopia, 2018 (n=422)

**Table 7:** Univariate and Multivariable analysis of factors associated with food consumption score among pregnant women attending ANC service at Shegaw Motta Hospital, Northwest Ethiopia, 2018 (n=422)

| **<u>Food Consumption Score</u>** |  |  |  |  |  |
| --- | --- | --- | --- | --- | --- |
| **Variables** | **Unacceptable** | **Acceptable** | **COR(95%CI)** | **AOR (95%CI)** | **P-value** |
| **Knowledgeable** | 51 | 147 | 1.00 | 1.00*** |  |
| **Not knowledgeable** | 30 | 196 | 2.244(1.362, 3.696) | 1.683(0.861, 3.288) | 0.128 |
| **Wealth Index Richest** | 18 | 47 | 1.00 | 1.00*** |  |
| **Rich** | 20 | 87 | 1.666(0.418, 1.880) | 2.244(0.819, 6.142) | 0.116 |
| **Middle** | 13 | 48 | 1.414(0.724, 3.008) | 1.463(0.622, 3.440) | 0.383 |
| **Poorer** | 11 | 36 | 1.253(0.561, 2.799) | 1.433(0.513, 4.002) | 0.492 |
| **Poorest** | 7 | 56 | 3.064(1.412, 7.147) | 3.258(1.221, 8.698) | 0.18 |
| **G. Age <=12 weeks** | 3 | 10 | 0.992(0.263, 3.745) | 1.355(0.215, 8.518) | 0.746 |
| **12-28 weeks** | 28 | 163 | 1.733(1.040, 2.886) | 1.681(0.925, 3.054) | 0.088 |
| **>=29 weeks** | 50 | 168 | 1.00 | 1.00*** |  |
| **Still birth Yes** | 9 | 23 | 0.579(0.257, 1.303) | 1.722(0.619, 4.794) | 0.298 |
| **no** | 72 | 318 | 1.00 | 1.00 |  |
| **Abortion Yes** | 12 | 23 | 0.416(0.197, 0.876) | 0.461(0.176, 1.211) | 0.116 |
| **No** | 69 | 318 | 1.00 | 1.00*** |  |

**Note:** COR, Crude odds ratio; AOR, Adjusted odds ratio; CI, Confidence interval;
\*\* Statistically significant
Others\*: include Muslim, Protestant and Catholic religion followers
1.00\*\*\* reference category

### 4.5. Factors associated with Food Consumption Score

In the univariat analysis, twelve independent variables, namely; study participants’ age, education status, occupation, residence, religion, knowledge, history of abortion, history of still birth, husband’s employment and occupation status, gestational age, and wealth status were found with a p-value of <0.25. However, at 5% level of significance multivariable binary logistic regression model analysis; only study participants’ residence and religion were significantly and independently associated with food consumption score.

The odds of unacceptable (poor/borderline) food consumption score were decreased by 92.7% (AOR =0.073 (95% CI :(0.021, 0.254)) in pregnant women who were Muslim, Catholic and Protestant religion followers as compared to those pregnant women who were Orthodox Christian followers.

Study participants who were living in the rural area were 4.59 times more likely to have unacceptable (poor/borderline) food consumption score as compared to those who were living in the urban area (AOR=4.594, 95% CI: 1.871, 11.283). Details of factors associated with food consumption score among pregnant women are presented in **Table7**.

## 5. DISCUSSION

Unacceptable food consumption score is the major public health problem (15) so strengthening nutrition intervention is essential (12). The current magnitude of Unacceptable food consumption score (poor or borderline) in this study was found to be 18.5% lied at the range (95% CI: 14.9, 22.5).

The magnitude of unacceptable (poor or borderline) food consumption score in the present study is considerably higher than reports from some developing countries, for instance in Armenia (11%), Bangladish (10%), Bennin (4%), Kenya (14%) and Gambia (3 %) (18-19). The less magnitude of unacceptable food consumption score in Armenia, Bangladish, Bennin and Gambia could be attributed to the better consumption of nutrient dense food and eating mixed diets. In addition, all study participants in Armenia were well educated, so that this may help them prevent to have unacceptable food consumption score. The previous national studies reported that food consumption score coverage in Ethiopia was 74 % (20) which is far lower than recommended by WFP which is 90% but this study finding is 81.5%; it may be due to that the national data covers large area which might be productive or resource scarce. However, the study area is very surplus, different nongovernmental organizations (NGOs) are working on pregnant women’s nutrition and the hospital has functional women nutrition committee. Since the study was at institutional level, pregnant women may have gotten knowledge about proper food consumption during ANC visit. The above listed possible points could explain the discrepancy.

The current study is lower than studies conducted in Nepal (34%), Uganda (30%) and Ethiopia (26%) of unacceptable food consumption score. The possible justification for this variation could be firstly, residence of the respondents have crucial role that is in Nepal only 8% of households having unacceptable food consumption who were living in urban area and 24% in the rural areas(15); Secondly, this might be related with poor food consumption utilization and mal distribution of food. For instance, food consumption utilization in rural Nepal was 66%)(15) which is lower than food consumption coverage in Ethiopia 74% **(20)**.

The possible discrepancy for this study could be, participants who were enrolled in the study were small in number which results in difficulty of generalizability, gestational age wealth status, geographical difference and methodological variation.

The present study showed significantly higher magnitude of unacceptable food consumption score as compared to previous studies conducted in Ethiopia; Amhara (11%), Tigray (16%), Afar (6%), and Somali (17%) (20) but, still the figure is unacceptably high. This could be poor utilization of food, poor dietary diversity practice and consumption of cereal with pulses is common in these areas. Another explanation could be decreasing in public awareness towards proper food utilization from time to time and ecological difference between two study settings could describe this discrepancy. Besides, availability issue, cultural issue and seasonal variation may explain the difference.

The current study came up with the evidence that respondent’s residence and religion were factors in the food consumption score during pregnancy. Pregnant women who were living in rural areas were highly affected by unacceptable food consumption score than pregnant women who were living in urban areas. It has been observed that pregnant women who were living in rural area were about 4.6 times more likely to have an unacceptable food consumption score as compared to those who were living in urban areas. This may be due to less awareness and less food diversification habits in rural areas. This is supported by a study from Nigeria, Uganda and Ethiopia in which to be rural is responsible factor for the development of unacceptable food consumption score (15, 21-22). It could also explained by the different level of awareness among two groups of population and cost of accessing better nutrition like; diary product and other diets which are enrich with micronutrients.

Those pregnant women who were Muslim, Protestant and Catholic followers were nearly 92.7% less likely to have unacceptable food consumption score as compared to those who were Orthodox Christian followers. This is supported by a review on religious recommendations on diet and life style all over the world and Ethiopian (23-24). The possible reason for this difference might be fasting and food preference.

## 6. LIMITATION

### 6.1. Limitation of the study

Social desirability bias

✓ Explaining the objectives
✓ Isolation

Recall bias and seasonality of the problem

✓ Used local events
✓ Probing

The Scientific value of the outcome may be affected due to not have done with ordinal logistic regression

## 7. CONCLUTIONS AND RECOMMENDATIONS

### 7.1. Conclusions

The magnitude of the unacceptable food consumption score among pregnant women attending ANC at Shegaw Motta Hospital was found to be high. Participants’ religion and residence were found to be statistically significant with a food consumption score during pregnancy.

### 7.2. Recommendations

#### Local decision makers

✓ Religion based nutrition education regarding food consumption score should be given for the community
✓ It would be better if special attention is given to pregnant women specially for those who are Orthodox Christian followers and living in rural areas

#### For researchers

∘ It will be better if the food consumption score will be done at community level
∘ Follow up study should be conducted to see the pregnancy outcomes
∘ Ordinal logistic regression is better to keep the scientific classification of the food consumption score

#### For pregnant women

∘ Maximize frequent intake of food items such as; meat, fish, milk etc.
∘ Diversified food intake in every day meal

## Data Availability

The data is not available

## ACKNOWLEDGEMENT

We would like to appreciate Shegaw Motta Hospital and study participants who were voluntary to participate in the study.

## Authors’ contribution

- MB-data collection and final draft writings, TW-edition, corrected and manuscript preparation, MT and ZA advise and comment. All authors approved the final manuscript.

## Declaration

- The authors declare that no competing interests.

